# What Your bowel Sounds Can Tell: The Hidden Language of Digestive Health

**DOI:** 10.64898/2026.03.15.26348419

**Authors:** Zahra Mansour, Verena Nicole Uslar, Dirk Weyhe, Thorsten Aumann-Münch, Danilo Hollosi, Nils Strodthoff

**Author notes:** Contributing authors.

## Abstract

**Purpose:** While bowel sound auscultation represents a key component of abdominal examination, its utility is limited because bowel sounds (BS) are intermittent, variable, and influenced by factors such as diet and digestive state. This renders it challenging to use them for a quantitative assessment of gastrointestinal health.

**Methods:** BS signals were recorded from 84 subjects (39 patients and 45 healthy controls) using an acoustic SonicGuard sensor and categorized into four patterns. Metadata on physiological parameters were collected to examine their influence on BS characteristics and the differences between healthy and patient BS patterns.

**Results:** Bowel sound patterns are significantly influenced by meal timing, caffeine consumption, and medication intake. Significant differences between healthy and patient groups were also observed in sound count, duration, energy, and waveform shape. These differences were mirrored in the performance of machine learning models finetuned for BS patterns classification, with performance depending on the group used for training and evaluation.

**Conclusion:** BS patterns present a promising quantitative indicators of gastrointestinal health when analyzed alongside relevant physiological parameters.

## 1 Introduction

### Abdominal auscultation as a diagnostic tool

The first systematic use of bowel sounds (BS) for diagnostic purposes was reported by Walter Cannon in 1905 [1], who established a link between bowel sound activity and underlying gastrointestinal physiology. Since then, BS auscultation has remained a fundamental component of abdominal examination. Traditionally, clinicians listen to BS for approximately two minutes, moving the stethoscope across several abdominal quadrants to determine whether BS are present or absent. Based on their occurrence and intensity, BS are typically classified as normal, hypoactive, or hyperactive[2]. However, no quantitative thresholds clearly distinguish these categories, and clinical assessment remains largely subjective. Although some guidelines recommend auscultating for up to five minutes before concluding that bowel sounds are absent based on the examiner’s subjective estimation of bowel activity [3], in practice, the actual listening times are often much shorter. Surveys have shown that only a minority of clinicians routinely auscultate all four quadrants, and many spend less than one minute per patient [4]. This variability further limits the diagnostic value of traditional BS auscultation. As a result, the outcome of auscultation depends heavily on the clinician’s ability to recognize BS occurrences, which is a subjective process influenced by individual experience. Studies indicate that approximately half of the clinicians are rarely able to recognize the BS occurrence, and only 17% reported that they are consistently able to define the BS[4]. These limitations highlight the need for automated detection and feature extraction of BS, enabling objective, quantitative, and reproducible assessment beyond the constraints of human hearing.

### The relationship between bowel sounds and digestive and physiological states

BS are primarily generated by muscular contractions of the intestines that propel food, liquids, and gas through the digestive tract, producing gurgling and rumbling noises as the hollow intestinal walls resonate [5]. Consequently, variations in digestive activity are indirectly mirrored in bowel sound characteristics. It is therefore reasonable to assume that BS patterns are influenced by factors such as food intake and the digestive stage at which auscultation occurs.

### Food intake

Several studies have investigated this relationship by recording BS before and after meals. For example, Vasseur et al. [6] found that the number, total duration, and mean energy of bowel sounds measured over ten-minute intervals increased significantly after food ingestion. Similarly, Sakata et al. [7] observed marked changes in bowel sound frequency following meals after prolonged fasting.

### Caffeine intake

Other studies have identified additional physiological factors that can modulate bowel sound characteristics. Caffeine intake, for instance, may increase gastrointestinal gas and stimulate motility, thereby influencing bowel sound length and frequency [8, 9].

### Medication intake

Likewise, medications that affect intestinal motility have been reported to either enhance or suppress bowel sound activity [**?**]. These findings underscore the importance of not only recording bowel sounds but also documenting the physiological conditions present during auscultation. Collecting such contextual information enables a more reliable interpretation of BS signals and allows the extracted features to better reflect the underlying gastrointestinal state.

### The relationship between BS and gastrointestinal diseases

Beyond the presence or absence of BS, certain pathological sound patterns are associated with specific gastrointestinal (GI) conditions. For instance, patients with mechanical obstruction often exhibit a characteristic pattern of clustered bowel sounds — three to ten regular sounds occurring approximately every five seconds — followed by a period of silence [4]. However, such pathological patterns are relatively rare, and it remains important to investigate whether variations in normal BS activity can indicate underlying health conditions. Clinical studies have shown that only about one-third of clinicians can recognize postoperative ileus based solely on auscultation, with affected patients exhibiting a significantly reduced number of bowel sounds compared with healthy controls [10]. In contrast, auscultation alone has proven insufficient to discriminate patients with bowel obstruction from healthy individuals [11]. More recently, research supported by advanced signal processing and machine learning methods has demonstrated the potential of automated BS in disease detection. For example, acoustic features derived from BS have been used to distinguish patients with inflammatory bowel disease (IBD) from healthy controls with promising accuracy [12–14]. These findings suggest that bowel sounds carry valuable physiological information reflecting gastrointestinal motility and pathology. Despite these advances, no comprehensive study has yet compared detailed acoustic characteristics patient and healthy populations to determine whether statistically significant differences exist.

### Research Questions

In this study we will take a first step toward understanding of the quantitative information content within the bowel sound signal. In particular, we aim to characterize (1) how bowel sounds are affected by the gastrointestinal health status of the patient (2) how bowel sound are influenced by different digestive and physiological states.

## 2 Methodology

One of the main challenges in BS research is the lack of comprehensive datasets that include both healthy and patient recordings, along with contextual information describing the physiological and digestive state of each subject during auscultation. To address this gap, the first step of this study involved the acquisition of BS recordings together with relevant physiological and digestive condition data. A second challenge lies in preparing these data for quantitative analysis. This requires not only detecting and labeling bowel sounds but also classifying them into distinct BS patterns. Once annotated, the relationship between the occurrence of these BS patterns and the corresponding physiological or digestive conditions can be analyzed. Finally, statistical tests can be applied to identify significant differences between healthy and patient groups in terms of BS characteristics. The overall methodology adopted in this study is summarized in Fig. 1 and is described in detail in the following sections.

**Fig. 1.**
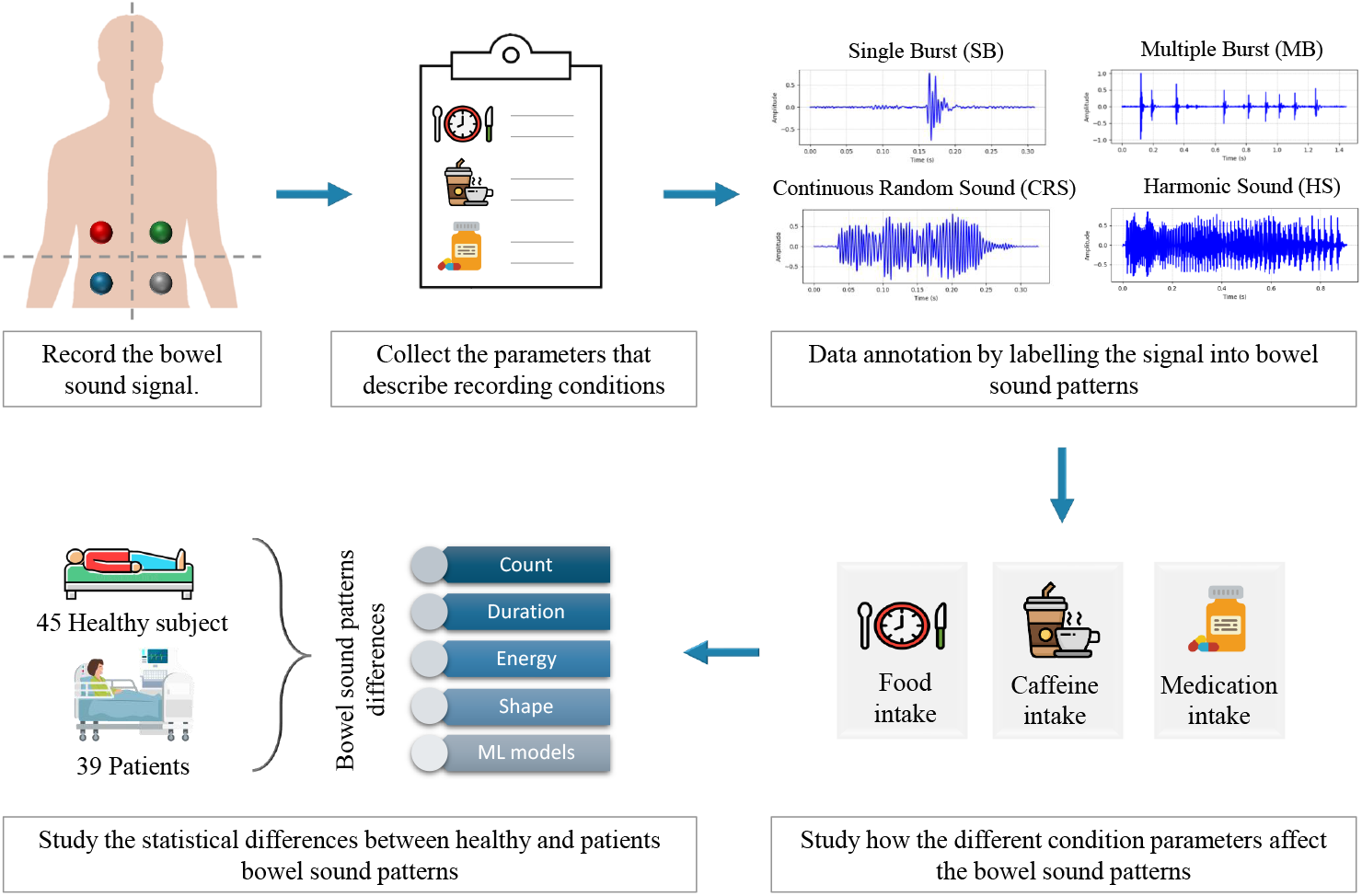
Overview of the study methodology. The process includes (1) acquisition of bowel sound (BS) recordings together with (2) physiological and digestive condition data, (3) annotation and classification of BS into distinct sound patterns (Single Burst (SB), Multiple Burst (MB), Continuous Random Sound (CRS), Harmonic Sound (HS)), (4) analysis of the relationship between BS pattern occurrence and physiological parameters, and (5) statistical comparison between healthy and patient groups to identify significant differences in BS characteristics.

### 2.1 BS Recording

#### Recording protocol

The research protocol underlying the data recordings was approved by the Medical Ethics Committee of the University of Oldenburg (2022-056). Before participation, subjects were informed about the purpose of the study and consented to the use of the collected data in an anonymized form. During the recording session, each subject was asked to lie down in a supine position. The BS were then recorded from the four abdominal quadrants (right upper quadrant (RUQ), left upper quadrant (LUQ), right lower quadrant (RLQ), left lower quadrant (LLQ)) as shown in Fig 1, using the SonicGuard wearable multi-channel acoustic sensor [15]. Seven minutes of BS were recorded from each quadrant with a total of 28 minutes of recording for each subject.

#### Participant information

A total of more than 40 hours of bowel sound recordings were acquired from 84 subjects, including 36 patients diagnosed with gastrointestinal (GI) diseases and 48 healthy controls, at PIUS Hospital in Oldenburg, Germany. Participants ranged in age from 20 to 83 years, comprising 40 females and 44 males, with body mass index (BMI) between 18 and 40 kg/m^2^. The patient group included individuals with colon cancer, Crohn’s disease, and postoperative gastrointestinal disorders.

### 2.2 Collection of digestive and physiological state parameters

Each participant answer a questionnaire right before the data recording, which provide basic information related to their recent food intake, caffeine consumption, and use of digestive-related medication. Based on these responses, subjects were grouped according to the time since their last meal, whether they had consumed caffeine shortly before the recording, and whether any medication taken within the past 24 hours was expected to influence digestive activity. This categorization allowed studying the differences in digestive and physiological states across participants during signal acquisition

### 2.3 Data annotation and bowel sound patterns

The BS patterns analyzed in this study were defined in detail in our previous work [16]. They align with classifications commonly used in abdominal auscultation and prior literature, covering all sound events observed in the dataset. The four bowel sound patterns are shown in Fig. 1, and represent distinct types of acoustic activity that differ primarily with respect to the anatomical location of sound generation within the gastrointestinal tract and the composition of the intestinal contents. Short, discrete sound events are typically associated with localized intestinal movements and fluid-dominated content, giving rise to clicking-like patterns such as **Single Burst (SB)** events (10–30 ms) and grouped **Multiple Burst (MB)** events (40–1500 ms total duration).

In contrast, sounds generated in the presence of larger amounts of air or during the movement of mixed contents through wider intestinal regions, such as the colon, tend to be more prolonged and resonant. These mechanisms are reflected in **Continuous Random Sound (CRS)** patterns (200–4000 ms), characterized by sustained bubbling or rumbling activity, and in **Harmonic Sound (HS)** patterns (50–1500 ms), which exhibit more pronounced resonance and echo-like components. Together, these four patterns provide a general categorization of bowel sounds based on differences in location, content, and physical sound propagation rather than specific pathological interpretations.

The recordings from 40 subjects (18 patients and 22 healthy controls) were manually annotated into the 4 BS patterns described above by a trained member of the medical staff using the Audacity software, based on both the waveform and spectrogram characteristics as well as auditory evaluation. The remaining 43 recordings were automatically annotated using the auto-annotation algorithm.The algorithm performs BS event detection followed BS classification and yields accurate BS event markers without human intervention[17].

### 2.4 The relationship between bowel sounds and digestive and physiological states

For each patient, we quantified bowel-sound (BS) activity by counting occurrences of each BS pattern and calculating a motility rate (occurrences per minute). Patients were grouped according to three physiological factors—food intake, caffeine intake, and medication use for more details see Appendix.1. For each factor, we compared the per-patient, per-minute frequency of each BS pattern across groups.

### 2.5 The relationship between BS and gastrointestinal diseases

Recordings were divided into Healthy and Patient groups for comparative analysis. For each bowel-sound event, duration, energy, and event counts were computed using the equations described in Appendix.3. Between-group differences in these features were assessed using the Mann–Whitney U test described in the Appendix.2.To evaluate whether the shape of the sound waves differed, we used Dynamic Time Warping to compare signal patterns described in the Appendix.4. Lastly, we tested whether these group differences affected machine-learning performance by fine-tuning three Audio Spectrogram Transformer (AST) models—one trained only on healthy data, one on patient data, and one on mixed data—and evaluating each on healthy, patient, and mixed test sets for more details check Appendix.5.

## 3 Results and discussion

### 3.1 Relationship between bowel sounds and digestive and physiological states

#### 3.1.1 Food intake

Subjects were categorized into four groups according to time since the last meal. Changes in the occurrence rate (events/min/subject) of each BS pattern, and of overall BS activity, across these meal–timing groups are shown in Fig. 2. A noticeable peak in the occurrence of MB and CRS patterns—along with an increase in overall BS activity—was observed at approximately 1 hour after food intake. This rise is consistent with the early phases of postprandial motility [18], when more complex digestive processes are initiated. In contrast, the HS pattern, which reflects stenosis rather than a physiological digestive phase, showed no appreciable change across the four timing groups (less than 0.01%), suggesting limited direct coupling to postprandial state.

**Fig. 2.**
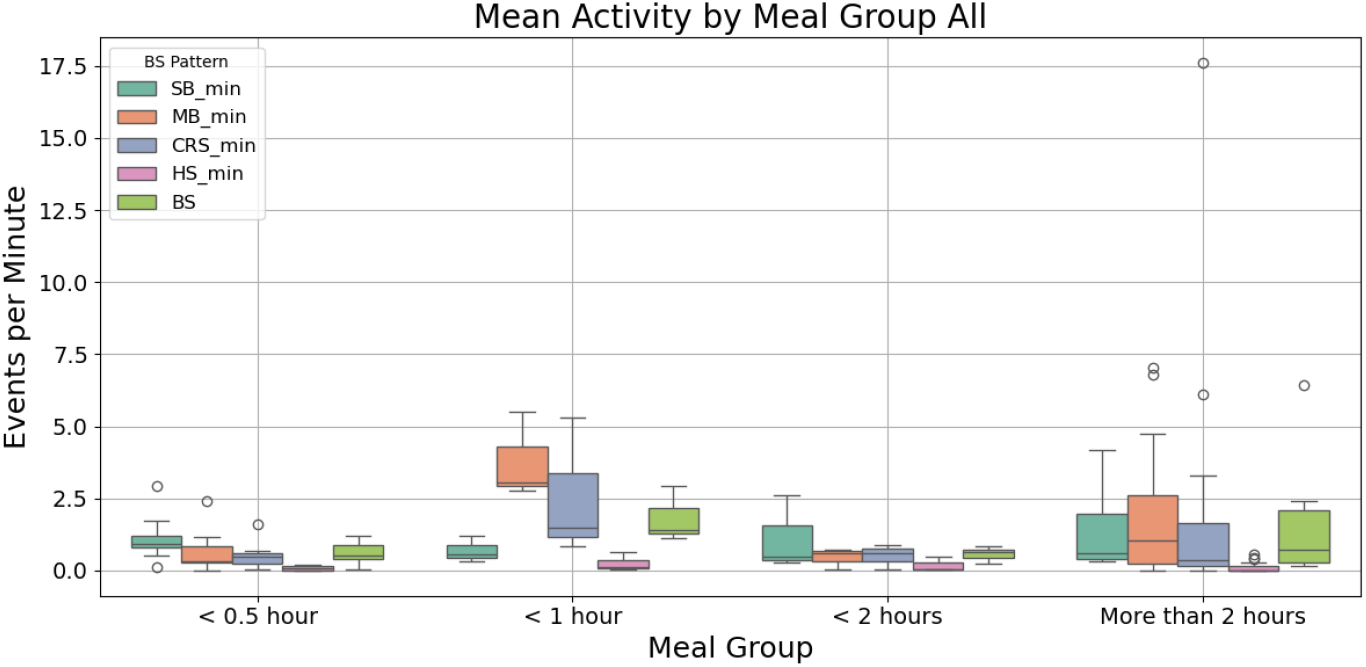
This boxplot show the changes in the occurrence rate of BS patterns per min per subject in respect to the time since last meal. A peak in MB and CRS (and overall BS activity) is seen at 1 h postprandial, whereas HS remains unchanged across groups. The *>*2 h bin spans a broad interval (3–12 h), which may blur timing effects.

Interpretation of the *>*2 hours category is more challenging because it spans a broad interval (*∼* 3–12 hours postprandial). Despite this heterogeneity, the data indicate a gradual increase in all BS patterns a few hours after food intake, although the wide window likely attenuates temporal resolution compared with narrower postprandial bins.

#### 3.1.2 Caffeine intake

Subjects were divided into two groups according to whether a caffeinated drink was consumed within the 30 minutes preceding the recording or not. For each group, the occurrence rate (events/min/subject) of each BS pattern and overall BS activity was computed and is shown in Fig. 3.

**Fig. 3.**
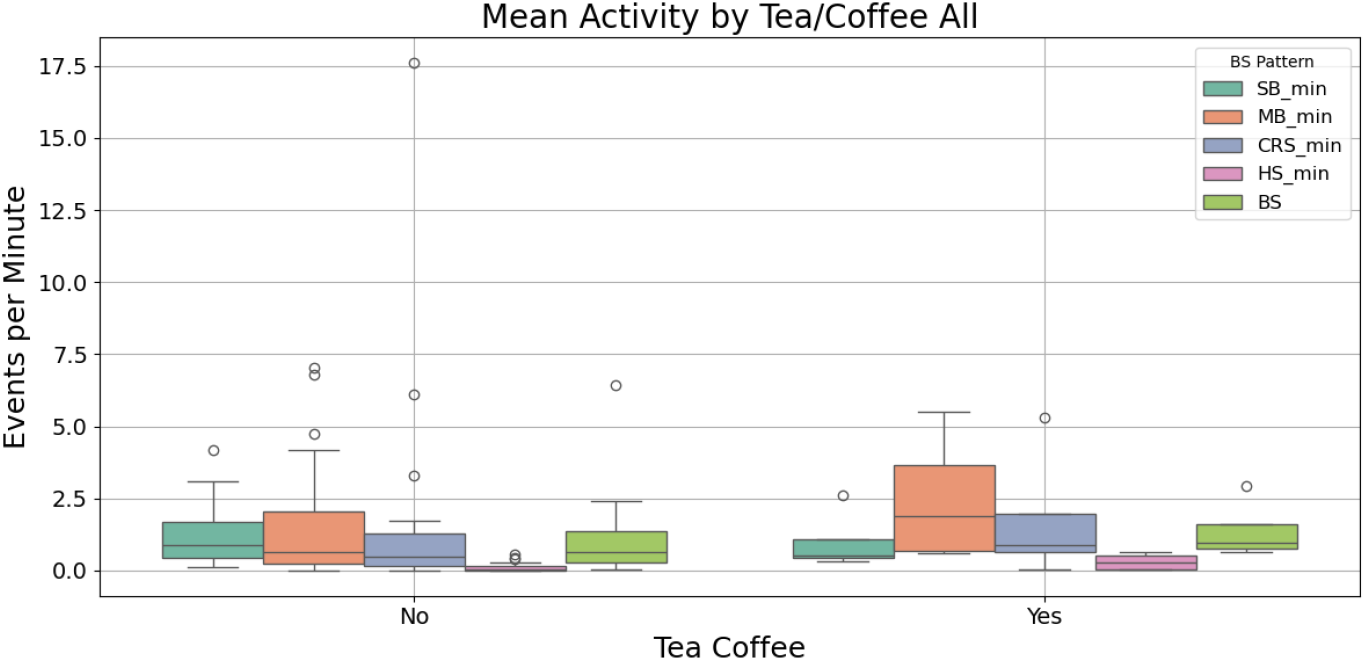
Occurrence rate (events/min/subject) of BS patterns with vs. without caffeine intake, increased rates are observed across patterns and overall BS activity except for SB, consistent with caffeine’s acute stimulatory effect on gastrointestinal motility.

Recordings obtained after recent caffeine intake exhibited higher occurrence rates for all BS patterns (except SB) as well as an overall increase in BS activity. This pattern is consistent with the acute stimulatory effects of caffeine on the nervous system and gastrointestinal motility, which may be reflected in enhanced BS activity[8, 9].

#### 3.1.3 Medication intake

Subjects were divided into three groups based on self-reported medication use within the 24 hours prior to recording: (i) no medication or medication with no expected gastrointestinal (GI) effect, (ii) medications expected to enhance GI motility, and (iii) medications expected to inhibit GI motility. Changes in the occurrence rate (events/min/subject) of each BS pattern and overall BS activity across these groups are shown in Fig. 4.

**Fig. 4.**
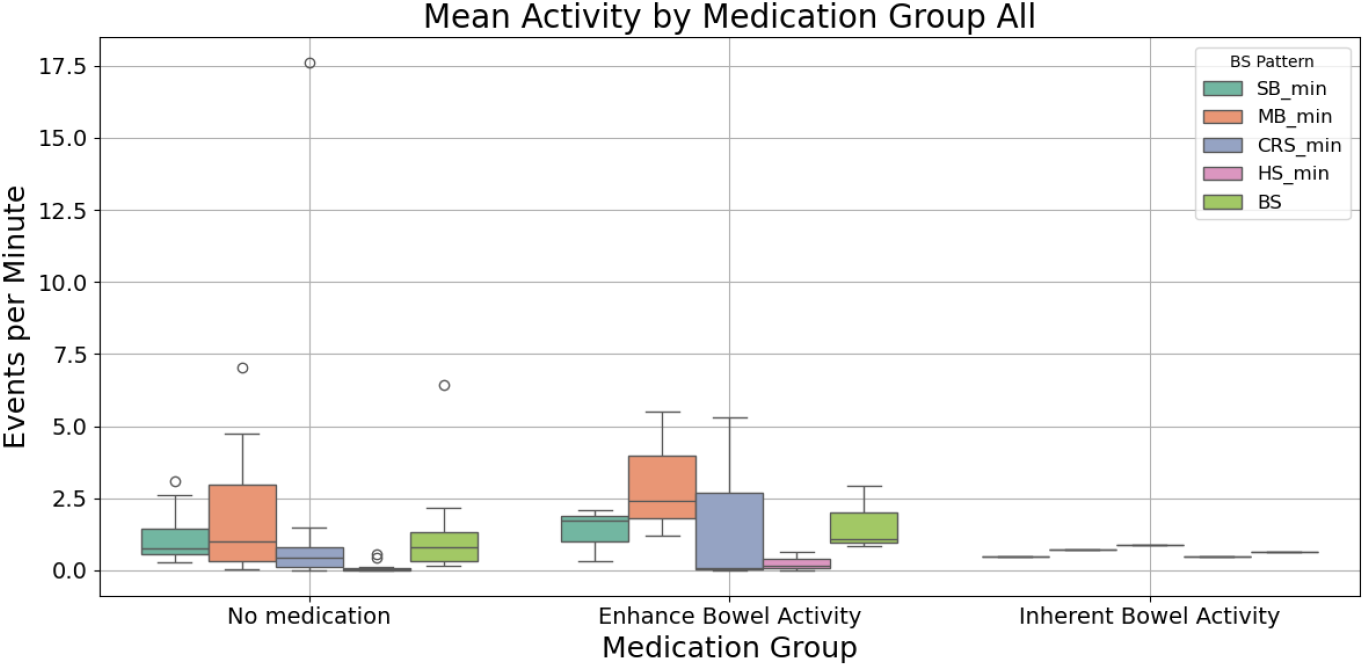
The occurrence rate (events/min/subject) of BS patterns by medication group within the preceding 24 h. Rates increase in the motility-enhancing group and decrease in the motility-inhibiting group, relative to the no–GI-effect group.

Compared with the no–GI-effect group, the motility-enhancing group exhibited a significant increase in the occurrence of all BS patterns as well as overall BS activity, consistent with a prokinetic effect[**?**]. In contrast, the motility-inhibiting group showed a marked reduction in BS activity across patterns, indicating suppression of bowel sounds. These findings suggest that BS monitoring may provide complementary, noninvasive insight into patient specific responses to medications that modulate GI motility.

### 3.2 Relationship between BS and gastrointestinal diseases

#### 3.2.1 BS features (Count, Duration and Energy)

To compare bowel sounds between healthy individuals and patients, all recordings were assigned to two groups (Healthy vs. Patient). For each recording, we extracted several acoustic features (Count, Duration and Energy) and analysed them at the subject level, stratified by bowel-sound pattern (SB, MB, CRS, HS). Group differences were evaluated using FDR–adjusted *p*-values, and effect sizes were quantified using Cliff’s *δ*. A summary of these statistical results for all bowel-sound features is presented in Table 1. Overall, *count* is consistently higher in Patients (large effects across SB, MB, CRS, and HS), indicating a greater frequency of bowel-sound (BS) events in the patient cohort. In contrast, *duration* tends to be higher in Healthy subjects for all labels, with effect sizes ranging from small (MB, CRS) to medium (SB) and large (HS), suggesting that individual events are typically longer in Healthy recordings. *Energy* shows smaller group contrasts (around 10^*−*7^): differences are negligible to small for SB and MB, small for CRS, and reaches a medium effect for HS, with healthy controls generally exhibiting higher per–event energy. All reported differences remain statistically significant (*p <* 0.05) after FDR control.

**Table 1.**
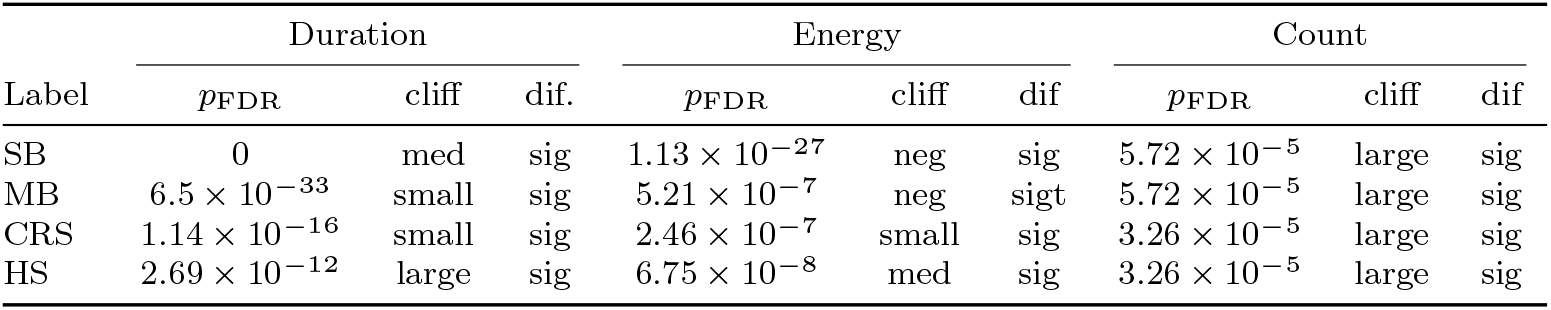
Per-label Mann–Whitney U results for Duration, Energy, and Count. Cells report FDR-adjusted *p*-values (*p*_FDR_), Cliff’s *δ* magnitude category (cliff: neg = negligible; small; med;large), and the significance flag (dif). By convention, *δ >* 0 indicates larger values in Healthy and *δ <* 0 in Patient.

**Table 1.** Physiological and digestive parameters collected from participants and their corresponding grouping criteria.

| Parameter | Categories / Groups |
| --- | --- |
| **Food intake**<br>(When you had your last meal) | • <30 min (12 subjects)<br>• <1 hour (15 subjects)<br>• <2 hours (14 subjects)<br>• More than 2 hours (43 subjects) |
| **Caffeine consumption**<br>(Did you drink coffee in the last 30 minutes?) | • Yes (37 subjects)<br>• No (47 subjects) |
| **Digestive medication intake**<br>(Did you take any medication in the last 24 hours?) | • Enhance digestion (7 subjects)<br>• Inherent digestion (7 subjects)<br>• No digestion effect or No medication (70 subjects) |

*Note:* Medication groups are defined as follows; **Enhance digestion:** prokinetic or enzyme-stimulating drugs; **Inherent digestion:** antispasmodic or motility-inhibiting medications; **No digestion effect or No medication:** drugs unrelated to gastrointestinal function.

Viewed per label within this overall pattern, members of the patient group produce more events of each type (large effects on Count), while Healthy recordings exhibit longer events (Duration) across SB, MB, CRS, and HS, and modestly higher Energy, most pronounced for HS. This configuration—higher event frequency in Patients coupled with shorter, lower–energy events—may reflect altered motility dynamics (e.g., dysrhythmic or fragmented activity) that increase the number of detectable transients without proportionally increasing their per–event duration or power. Conversely, longer and somewhat more energetic events in Healthy subjects are consistent with more organized contractions and clearer acoustic signatures. Together, these findings indicate that pathology is associated primarily with an increase in event frequency, whereas event morphology (duration and energy) remains equal or shifts in favor of Healthy recordings, with label–specific magnitudes detailed in Table 1.

#### 3.2.2 BS shape differences using DTW

While earlier analyses showed some variation in features such as the Energy of the BS between healthy and Patient signal, they did not address whether the underlying waveform morphology differed. To answer this question, Dynamic Time Warping (DTW) was performed by measuring the distances between Mel-cepstral sequences, and revealed systematic shape differences between Healthy and Patient recordings (Table 2). For all labels (SB, MB, CRS, HS), the between–group distances were significantly larger than the pooled within–group distances, indicating that acoustic shapes from Patient recordings differ from those of Healthy controls beyond the variability observed within each cohort.

**Table 2.** DTW mean distances within Healthy (within H), within Patient (within P), and between groups (H&P) for each BS label. Larger between-group distances (with MWU *p*-values) indicate shape differences; “shapes differ” marks significant contrasts.

| Label | mean dist | | | $p$ | shape differ |
| --- | --- | --- | --- | --- | --- |
|  | within H | within P | between H&P |  |  |
| SB | 4139.5 | 890.2 | 2796.2 | $1.15 \times 10^{-5}$ | shapes differ |
| MB | 25785.0 | 14255.8 | 21830.9 | $3.72 \times 10^{-5}$ | shapes differ |
| CRS | 41344.1 | 13597.1 | 27109.7 | $1.117 \times 10^{-4}$ | shapes differ |
| HS | 27418.6 | 7363.1 | 21692.8 | $6.81 \times 10^{-28}$ | shapes differ |

A consistent pattern emerged across labels: within–Patient distances were the smallest, within–Healthy distances were the largest, and between–group distances lay in between (e.g., MB: within H = 25,785, within P = 14,256, between = 21,831). This configuration suggests comparatively greater shape homogeneity within the Patient cohort and greater heterogeneity within the Healthy cohort, with an additional mismatch introduced when aligning across cohorts. The separation was most pronounced for HS (within H = 27,419; within P = 7,363; between = 21,693; *p* = 6.81 × 10^*−*28^), consistent with HS capturing morphological characteristics that diverge strongly across cohorts. MB and CRS showed the same qualitative pattern with significant, intermediate separation (MB *p* = 3.72 × 10^*−*5^; CRS *p* = 1.12 × 10^*−*4^), and SB exhibited a smaller but still significant effect (*p* = 1.15 × 10^*−*5^).

#### 3.2.3 Data-driven perspective on Healthy vs Patient cohorts

To investigate whether the underlying differences between bowel sound patterns of healthy individuals and patients are reflected in the performance of the classification model, we evaluated the model under several training and testing configurations. Table 3 summarizes one-vs-all performance under three training regimes (Healthy-only, Patient-only, All) and three test sets (Healthy, Patient, Mixed). A clear cohort effect is evident. Models trained and tested within the same cohort achieve the strongest results (Healthy *→*Healthy: avg = 0.94; Patient*→* Patient: avg = 0.99), indicating strong within-domain separability of BS classes. In contrast, cross-cohort generalization is asymmetric: the Patient-trained model transfers poorly to Healthy data (avg = 0.62; notably low for HS = 0.42), whereas the Healthy-trained model remains relatively stable on Patient data (avg = 0.93). Training on the combined cohort yields robust, near-ceiling performance on Patient data (avg = 0.96) and balanced results on Healthy and Mixed sets (avg = 0.68 and 0.95, respectively), suggesting improved resilience to domain shift.

**Table 3.** Train–test AUC (one-vs-all) for models trained on Healthy, Patient, or All subjects and evaluated on Healthy, Patient, and Mixed test sets; bold indicates the best performance.

| Train | Test | None vs all | SB vs all | MB vs all | CRS vs all | HS vs all | avg |
| --- | --- | --- | --- | --- | --- | --- | --- |
| Healthy | Healthy | **0.96** | **0.95** | **0.89** | **0.92** | **0.92** | **0.94** |
|  | Patients | 0.94 | 0.96 | 0.92 | 0.91 | 0.89 | 0.93 |
|  | All | 0.94 | 0.96 | 0.92 | 0.92 | 0.89 | 0.93 |
| Patients | Healthy | 0.70 | 0.64 | 0.68 | 0.65 | 0.42 | 0.62 |
|  | Patients | **0.99** | **0.99** | **0.99** | **0.99** | **0.99** | **0.99** |
|  | All | 0.98 | 0.98 | 0.98 | 0.96 | 0.97 | 0.98 |
| All | Healthy | 0.78 | 0.68 | 0.75 | 0.67 | 0.51 | 0.68 |
|  | Patients | **0.97** | **0.99** | **0.94** | **0.95** | **0.95** | **0.96** |
|  | All | 0.96 | 0.97 | 0.94 | 0.94 | 0.93 | 0.95 |

Across BS patterns, within-cohort peaks (marked in bold face) show that each model best recognizes the distributions it was trained on. The pronounced drop of the Patient-trained model on Healthy HS likely reflects cohort-specific spectral–temporal signatures that do not generalize when pathology-related cues are absent. Conversely, the Healthy-trained model maintains high performance on Patient data, consistent with broader, less pathology-specific representations learned from non-diseased recordings. Overall, these findings show that Healthy–Patient differences extend beyond variations in simple acoustic features and are also evident at the structural level, indicating that the BS waveform itself differs between healthy and patient groups. These signal-level differences propagate to model behaviour: training on cohort-matched data yields the highest discrimination within that cohort, whereas training on mixed data provides a practical balance between achieving strong within-domain AUC and maintaining robustness across cohorts.

## 4 Limitations and Future Directions

This study takes further step toward deepening our understanding of the information embedded in BS signals and the physiological factors that may influence them. To provide more definitive answers to the research questions raised here, future work would benefit from a larger and more focused dataset. In particular, the patient cohort in this study included a heterogeneous mix of gastrointestinal conditions, such as inflammatory and ileus-related disorders. Expanding the number of recordings within specific disease groups would not only strengthen comparisons between healthy and patient cohorts, but also enable meaningful discrimination among different disease categories.

More standardized recording conditions would further enhance the interpret ability of BS features. In the current dataset, parameters such as time since last meal varied within groups, making it more challenging to isolate the effects of individual factors. Establishing more controlled conditions in future studies would help address this issue. Continued development of automated BS recording and detection tools will also play a key role in enabling high-quality, consistent data collection and supporting more refined analyses.

## 5 Conclusion

Traditional bowel-sound (BS) auscultation captures limited information. In contrast, automated BS detection with feature extraction (count, duration, energy, and temporal–spectral shape) reveals clinically relevant patterns. We observed a postprandial peak in BS activity at approximately 1 hour after food intake, a marked increase following recent caffeine consumption, and medication-related modulation consistent with expected pro- and anti-motility effects—supporting BS monitoring as a noninvasive marker of physiological state.

Across health status, patients showed higher event counts, whereas healthy recordings tended to have longer and more energetic events with distinct shapes. These differences likely reflect altered motility dynamics in disease (more frequent, shorter, lower-energy transients). The same cohort differences appeared at the model level: classifiers performed best when trained and tested within the same cohort, and mixed-cohort training improved robustness to domain shift.

Further validation with larger datasets and more controlled subgrouping (varying one factor at a time) is needed to strengthen generalizability and refine clinical thresholds. Overall, BS analysis offers a promising, scalable tool for monitoring digestion, probing medication effects, and distinguishing health states.

## Data Availability

All data produced in the present study are available upon reasonable request to the authors

https://figshare.com/articles/media/Bowel_sounds_signal/28595741/1

## Acknowledgements

The authors acknowledge intramural funding from Faculty VI, Carl von Ossiezky Universität Oldenburg (Forschungspool, Potentialbereich mHealth) and from MWK Niedersachsen (via Fraunhofer IDMT, institute part HSA, project Connected Health).

## Appendices

### .1 Physiological parameters

The main physiological parameters measured in this study and their corresponding grouping criteria are summarized in Table 1.

### .2 Mann–Whitney U test and effect size (Cliff’s *δ*)

Because the data used in this study are non-normally distributed with unequal variances, group comparisons were performed using the Mann–Whitney U (Wilcoxon rank–sum) test, a robust non-parametric procedure for two independent samples [19, 20]. The test ranks all observations jointly and evaluates whether one group tends to have higher ranks than the other (i.e., a shift in distributions).

The *p*-value for the Mann–Whitney test was computed as follows. Let groups have sizes *n*_*i*_ and *n*_*j*_ (*N* = *n*_*i*_ + *n*_*j*_) and pooled ranks with sum *R*_*i*_ for group *i*. The Mann–Whitney statistics are:

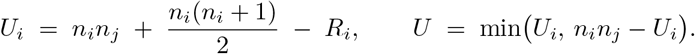

Under the null hypothesis of no difference between groups, *U* is approximately normally distributed with

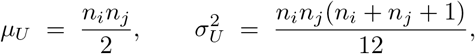

optionally adjusted for ties in the ranks. A standardized test statistic is then obtained as

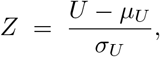

and the two-sided *p*-value is computed from the standard normal distribution as

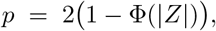

where Φ(.) denotes the cumulative distribution function of the standard normal distribution.

#### Effect size

Alongside *p*-values, **Cliff’s delta** (*δ*) was reported to quantify magnitude and direction of the difference [21]. It can be interpreted as

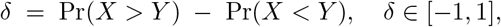

where *δ* = 0 indicates no dominance, *δ >* 0 that group *X* tends to have larger values, and *δ <* 0 the reverse. Conventional thresholds were used for interpretation: negligible (|*δ*| *<* 0.147), small (0.147 *≤* |*δ*| *<* 0.33), medium (0.33 *≤* |*δ*| *<* 0.474), and large (|*δ*| *≥* 0.474).

#### Statistical significance

Two-sided significance was assessed at *α* = 0.05, with control of the false discovery rate across families of tests via the Benjamini–Hochberg procedure; results were considered statistically significant when the FDR-adjusted *p <* 0.05 [22].

### .3 Features derived from bowel sound recordings

To compare bowel sounds between healthy individuals and patients, recordings were assigned to two independent groups (Healthy vs Patient). For each recording, the following features were derived and subsequently analysed at the subject level (stratified by BS pattern: SB, MB, CRS, HS).

#### Count (events/min/subject)

Event occurrence rate was computed as the number of BS events per minute for each pattern and subject (events/min/subject). Subject-level summaries (e.g., median per pattern) were used for between-group comparisons.

#### Duration (s)

Event duration was defined as the difference between the end and start timestamps of each BS event,

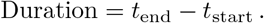

Per-subject, per-pattern distributions of event duration were obtained for inference.

#### Energy (dimensionless)

For each BS event, signal energy was calculated as the mean squared amplitude over the event window:

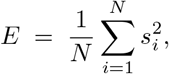

where *s*_*i*_ is the (unitless) signal amplitude at sample *i* and *N* is the number of samples within the event. Because amplitudes were scaled to *−*[1, 1], *E* is *dimensionless*. Events with missing or invalid audio were excluded. For comparability across subjects,energies were then standardised to mean zero and unit variance,

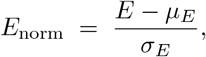

which is also dimensionless.

### .4 Temporal and spectral shape comparison using Dynamic Time Warping (DTW)

Potential differences in the temporal–spectral shape of bowel-sound (BS) events between Healthy and Patient groups were assessed using Dynamic Time Warping (DTW) applied to Mel-frequency cepstral coefficient (MFCC) representations [23]. For each event label (SB, MB, CRS, HS), up to 50 events per group were randomly sampled. Each BS segment was transformed into a sequence of 13-dimensional MFCC vectors, and DTW was used to compute a distance between pairs of MFCC sequences. For each label, three DTW distance distributions were obtained: within-Healthy, within-Patient, and between-group (Healthy vs Patient). To test whether Patient shapes systematically differed from Healthy shapes, a one-sided Mann–Whitney U (MWU) test compared the between-group distances against the pooled within-group distances.

For interpretability, the mean DTW distance was reported for each distribution (within-Healthy, within-Patient, between-group), together with the MWU statistic and its *p*-value. A shape difference was declared when *p <* 0.05.

### .5 Model-level reflection of Healthy vs Patient data differences

While the discussed features and measures represent widely used metrics to characterize differences between two sets of audio samples, they might miss essential features to characterize potential differences. We therefore complement these measures using a data-driven measure for dataset shift between different subsets, in line with the notion of black-box shift detection [24]. We use audio spectrogram transformer models finetuned on specific subsets as base models and consider the change in output probabilities when applied to different subset. To test whether acoustic differences between Healthy and Patient bowel-sound (BS) data persist at the model level, an Audio Spectrogram Transformer (AST) classifier was used [25]. AST applies a transformer to 2D log Mel-spectrograms by partitioning each spectrogram into non-overlapping patches that are embedded and processed via self-attention, enabling the capture of local spectral patterns and longer-range temporal dependencies. The model was initialised from weights pre-trained on the AudioSet corpus to provide generic audio representations [26].

Three cohort-specific AST models were fine-tuned: (i) Healthy-only, (ii) Patient-only, and (iii) a combined Healthy+Patient model. All used identical preprocessing (128-band log Mel-spectrograms; 25 ms window, 10 ms hop; per-band mean–variance normalisation), the same (train, validation, test) split (70%, 15%, 15%), and matched hyperparameters, so observed differences reflect cohort/domain effects rather than training variability.

Performance was summarized by the area under the receiver operating characteristic curve (AUC), a threshold-independent measure of discrimination. Each model was evaluated on three test sets (Healthy-only, Patient-only, Mixed) to assess within-domain AUC, cross-domain generalization, and robustness to cohort shifts.

## References

[1] Cannon, W.B.: Auscultation of the rhythmic sounds produced by the stomach and intestines. American Journal of Physiology-Legacy Content 14(4), 339–353 (1905)

[2] Li, B., Wang, J.-R., Ma, Y.-L.: Bowel sounds and monitoring gastrointestinal motility in critically ill patients. Clinical Nurse Specialist 26(1), 29–34 (2012) 10.1097/NUR.0b013e31823bfab8

[3] Bickley, L.S.: Bates’ Guide to Physical Examination and History Taking, 13th edn. Wolters Kluwer, Philadelphia, PA (2021)

[4] Felder, S., Margel, D., Murrell, Z., Fleshner, P.: Accuracy of abdominal auscultation for bowel obstruction. Journal of Surgical Research (2014) 10.1016/j.jsurg.2014.02.003

[5] Kölle, K., Aftab, M.F., Andersson, L.E., Fougner, A.L., Stavdahl, Ø.: Data driven filtering of bowel sounds using multivariate empirical mode decomposition. BioMedical Engineering OnLine 18(1), 28 (2019) 10.1186/s12938-019-0646-1

[6] Vasseur, C., Devroede, G., Dalle, D., Houtte, N., Bastin, E., Thibault, R.: Postprandial bowel sounds. Biomedical Engineering, IEEE Transactions on BME-22, 443–448 (1975) 10.1109/TBME.1975.324522

[7] Sakata, O., Matsuda, K., Suzuki, Y., Satake, T.: Basic study of occurrence frequency of bowel sounds after food ingestion. TENCON 2011 - IEEE Region 10 Conference, 1203–1206 (2011) 10.1109/TENCON.2011.6129303

[8] Horiyama, K., Emoto, T., Haraguchi, T., Uebanso, T., Naito, Y., Gyobu, T., Kanemoto, K., Inobe, J., Sano, A., Akutagawa, M., Takahashi, A.: Bowel sound-based features to investigate the effect of coffee and soda on gastrointestinal motility. Biomedical Signal Processing and Control 66, 102425 (2021) 10.1016/j.bspc.2021.102425

[9] Boekema, J., Samsom, M., Berge Henegouwen, G.P., Smout, A.J.P.M.: Coffee and gastrointestinal function: Facts and fiction: A review. Scandinavian Journal of Gastroenterology 34(230), 35–39 (1999) 10.1080/003655299750025525

[10] Felder, S., Margel, D., Murrell, Z., Fleshner, P.: Usefulness of bowel sound auscultation: A prospective evaluation. Journal of Surgical Education 71(5), 768–773 (2014) 10.1016/j.jsurg.2014.02.003

[11] Breum, B.M., Rud, B., Kirkegaard, T., Nordentoft, T.: Accuracy of abdominal auscultation for bowel obstruction. World Journal of Gastroenterology 21(34), 10018–10024 (2015) 10.3748/wjg.v21.i34.10018

[12] Tomomasa, T., Morikawa, A., Sandler, R.H., Kaneko, H., Masumoto, K., Kasai, T., Hyman, P.E.: Gastrointestinal sounds and migrating motor complex in fasted humans. American Journal of Gastroenterology 94(2), 374–381 (1999) 10.1111/j.1572-0241.1999.00816.x

[13] Shafik, A., El-Sibai, O., Ahmed, I., Shafik, I.A.: Bowel sounds: physiology and pathophysiology. Theoretical Biology and Medical Modelling 1(1), 6 (2004) 10.1186/1742-4682-1-6

[14] Niu, J., Hanai, T., Kudo, T., Takahashi, T., Yamada, Y., Nakamura, K., Tanaka, T., Sasaki, Y.: Development of a non-invasive bowel sound analysis system for evaluating gastrointestinal motility in patients with inflammatory bowel disease. PLOS ONE 13(12), 0208646 (2018) 10.1371/journal.pone.0208646

[15] Mansour, Z., Uslar, V., Weyhe, D., Hollosi, D., Strodthoff, N.: Sonicguard sensor—a multichannel acoustic sensor for long-term monitoring of abdominal sounds examined through a qualification study. Sensors 24(6), 1843 (2024) 10.3390/s24061843

[16] Mansour, Z., Uslar, V.N., Weyhe, D., Hollosi, D., Strodthoff, N.: Benchmarking machine learning for bowel sound pattern classification – from tabular features to pretrained models. PLOS ONE 21(1), 1–16 (2026) 10.1371/journal.pone.0338911

[17] Mansour, Z., Uslar, V., Weyhe, D., Hollosi, D., Strodthoff, N.: Towards Objective Gastrointestinal Auscultation: Automated Segmentation and Annotation of Bowel Sound Patterns (2026). 10.48550/arXiv.2603.07215. https://arxiv.org/abs/2603.07215

[18] Sakata, O., Suzuki, Y., Matsuda, K., Satake, T.: Temporal changes in occurrence frequency of bowel sounds both in fasting state and after eating. Journal of Artificial Organs 16(1), 83–90 (2013) 10.1007/s10047-012-0666-0. Epub 2012 Oct 30

[19] Mann, H.B., Whitney, D.R.: On a test of whether one of two random variables is stochastically larger than the other. Annals of Mathematical Statistics 18(1), 50–60 (1947)

[20] Wilcoxon, F.: Individual comparisons by ranking methods. Biometrics Bulletin 1(6), 80–83 (1945)

[21] Cliff, N.: Ordinal Methods for Behavioral Data Analysis. Psychology Press, ??? (1996)

[22] Benjamini, Y., Hochberg, Y.: Controlling the false discovery rate: A practical and powerful approach to multiple testing. Journal of the Royal Statistical Society: Series B 57(1), 289–300 (1995)

[23] Müller, M.: Information Retrieval for Music and Motion. Springer, ??? (2007)

[24] Lipton, Z.C., Wang, Y.-X., Smola, A.: Detecting and Correcting for Label Shift with Black Box Predictors (2018). 10.48550/arXiv.1802.03916. https://arxiv.org/abs/1802.03916

[25] Gong, Y., Chung, Y.-A., Glass, J.: Ast: Audio spectrogram transformer. In: Proc. Interspeech (2021)

[26] Gemmeke, J.F., al.: Audioset: An ontology and human-labeled dataset for audio events. In: Proc. ICASSP (2017)

